# Detection of TP53 mutations by IHC in acute myeloid leukemia varies with interpreter expertise and mutation status

**DOI:** 10.1101/2024.11.07.24316929

**Authors:** Lee P. Richman, Brianna Waller, Scott B. Lovitch, Ashwini Jambhekar

## Abstract

*TP53* mutations, including missense and inactivating (frameshift, splice site, and nonsense) mutations, occur in approximately 10% of myeloid neoplasms and confer adverse outcomes. Classification of myeloid neoplasms by both the World Health Organization and the International Consensus Classification standards now recognize the prognostic and therapeutic importance of early detection of *TP53* mutations. p53 immunohistochemistry (IHC) is a simple and rapid method commonly used to detect p53 mutations. More recently, sequencing via targeted panels has also seen increased use. While highly accurate, sequencing is resource intensive and not universally available. IHC represents a more accessible option for mutation detection, however previous studies have demonstrated variable accuracy, especially for inactivating *TP53* mutations. Using 134 bone marrow core samples of acute myeloid leukemia (AML) evaluated for *TP53* mutation by a sequencing panel, we assessed the concordance of p53 IHC with sequencing as well as the inter-rater reliability for IHC intensity and percent positivity. Consistent with previous studies, we found that p53 IHC was strongly specific and modestly sensitive for missense mutations, and that overall performance improved with dedicated hematopathology training. We also found that IHC performed poorly for inactivating mutations and was even variable between cases harboring identical amino acid changes. Low predicted transcriptional activity of *TP53* missense mutations correlated with a mutant pattern of IHC staining. The status of the second allele in missense mutations and variant allele fraction also affected the accuracy of p53 IHC as a surrogate for *TP53* allele status. AMLs expressing p53 mutations that were predicted to have low transcriptional activity correlated with reduced overall survival. Our results demonstrate limited practical utility of p53 immunohistochemistry for accurate evaluation of *TP53* mutation status due to multifactorial confounders.

## INTRODUCTION

Mutations in *TP53*, which encodes a tumor-suppressive transcription factor, occur in approximately 10% of myeloid neoplasms and confer an adverse prognosis with decreased treatment response, increased risk of relapse, and poorer overall survival.^1–5^ The most common alterations in *TP53* allele status, single nucleotide substitutions, are not detected by conventional cytogenetics and fluorescence *in situ* hybridization.^6–8^ Large targeted sequencing panels or, less commonly, dedicated *TP53* locus sequencing, are the gold standard for determining the presence of p53 mutations for appropriate classification of myeloid neoplasms in accordance with the World Health Organization and International Consensus Classification standards.^9^ However, such tests are resource-intense, batched with other samples to reduce waste, not universally available, and may be reported days to weeks after morphologic diagnosis.^5,9^ p53 overexpression or complete loss of protein expression frequently results from these mutations, therefore protein detection by p53 immunohistochemistry (IHC), which is widely available, has been proposed as a rapid surrogate for early detection of *TP53* variants in myeloid neoplasms. Previous studies focused on missense mutations and reported variable concordance with sequencing.^10–17^

*TP53* variants that confer adverse prognosis include missense mutations that reduce function (accounting for approximately 80% of cases with mutations) and inactivating “null” mutations arising from nonsense, frameshift, or splicing site mutations.^6–8^ Missense mutations, which are present most commonly in the DNA binding domain,^18^ can still have equivalent loss of function impact through a dominant-negative effect, as p53 operates as a tetramer.^7^ These missense mutations typically result in p53 accumulation due to their inability to induce the MDM2, the E3 ubiquitin ligase for p53,^19^ whereas null-type mutations commonly occur outside the DNA-binding domain result in loss of p53.^18^ These two states of abnormal protein accumulation and complete loss of expression are reflected by IHC as strong, overexpressed nuclear staining (graded as “3+ intensity”) and a complete absence of nuclear staining (graded as “0 intensity”), respectively.^10–13,20^ Previous work has demonstrated usefulness for minimal residual disease detection and has shown variable efficacy of p53 IHC for *TP53* mutation detection in myeloid neoplasms.^10–14^ Numerous confounding factors– such as sampling variability, percent of marrow involvement,^21,22^ high intensity staining in scattered erythroid precursors,^23^ variant allele fraction,^24^ and transcriptional activity of the specific mutant allele^25,26^ - may render the results of p53 IHC in bone marrow cores discordant with sequencing results. Previous studies have also demonstrated limited utility for null mutations and in the presence of specific additional cytogenetic abnormalities.^12,14–16,27,28^ An effective assay for detecting p53 mutations to triage patients at risk of adverse outcomes requires that it be robust to such confounding factors.

In addition to the functional impact of a missense mutation driving mutant protein accumulation compared to a null mutation causing total loss of expression, an increasing body of literature supports the assertion that each unique missense mutation has a varying degree of residual transcriptional activity, and thus, expression and function.^25^ Furthermore, the level of residual transcriptional activity of individual *TP53* missense mutations may be prognostic in myeloid neoplasms.^26^ In addition to the aforementioned sources of technical and genetic variability in the correlation between *TP53* allele status and the expected level of protein expression detectable by IHC, these findings suggest that the unique character of an individual mutation may influence the utility of IHC as a diagnostic approach. The effect of an individual mutation’s transcriptional activity on diagnostic p53 IHC has not been explored.

To determine the practical efficacy of p53 IHC for detecting null and missense *TP53* mutations, we evaluated the predictive performance of p53 IHC in 134 acute myeloid leukemia (AML) bone marrow core biopsies. We found modest performance that improved with dedicated hematopathology training, but was weakest with null mutations, consistent with previous studies,^12,14–16^. We identified multiple factors that impact accuracy of p53 IHC, including predicted residual transcriptional activity, second allele status, and variant allele fraction. Furthermore, the predicted residual transcriptional activity of p53 mutants correlated with overall survival. Overall, p53 IHC was specific but only modestly sensitive for detection of missense and null *TP53* mutations and sequencing should be prioritized for accurate diagnosis.

## METHODS

### Case Selection

Our case cohort included bone marrow core biopsies with archival Bouin’s-fixed paraffin embedded tissue from patients with a pathologic diagnosis of acute myeloid leukemia and concurrent high throughput sequencing molecular diagnostics that were treated at the Dana Farber Cancer Institute and Brigham and Women’s Hospital from 2017 to 2022. Cases included new and follow-up diagnoses. Paraffin blocks were retrieved and new unstained slides were prepared for p53 immunohistochemical staining.

Eighty biopsies with missense mutations in p53 were identified, and thirty-four wild-type (WT) cases were included. Twenty biopsies with null mutations (splice variant, frameshift, or nonsense mutations) were included. In the setting of multiple p53 variants, the case was classified by the variant with the highest variant allele fraction.

All studies were performed in accordance with a protocol approved by the Institutional Review Board of Brigham and Women’s Hospital.

### Histology and p53 immunohistochemistry (IHC)

Immunohistochemical studies were performed on 5-μm-thick Bouin’s-fixed, paraffin-embedded tissue sections using a BOND-III Immunostainer (Leica Biosystems). Epitope retrieval was performed with Epitope Retrieval 2 (Leica Biosystems #AR9640) on the BOND-III Immunostainer. The antibody used for p53 IHC was mouse monoclonal antibody DO-7 at a dilution of 1:100 (Agilent/Dako #M700101-2). The BOND Polymer Refine Detection system (3,3’-diaminobenzidine, brown chromogen, Leica Biosystems) was used for signal detection. Following the completion of immunohistochemical staining, the slides were briefly immersed in 1% copper sulfate solution, washed, briefly immersed in methyl green, washed, dehydrated, and cover-slipped.

### IHC intensity and percent positivity scoring

Three raters at different levels of training, an anatomic pathology resident, hematopathology fellow, and hematopathology attending (L.P.R., B.F.W., and S.B.L. respectively) reviewed all cases and scored IHC intensity as 0, 1+, 2+, and 3+. Staining was defined as:

- Absence of staining in blasts with intact background wild-type patchy positivity
- Weak staining in rare blasts, indistinguishable from background uninvolved bone marrow
- Moderate intensity nuclear staining
- Strong nuclear staining

The highest intensity staining comprising at least 5% of total cellularity was taken as the intensity score for the case. Percent positivity was determined by estimation of the percent of marrow cellularity composed of blasts with mutant (0,3+) or ambiguous (2+) staining on the slide.

The consensus staining for the case was defined as the score given by two or more raters. “No consensus” indicates no raters agreed and these cases were excluded from the indicated analyses.

### Molecular analysis

p53 mutation status was abstracted from the electronic medical record from concurrent high throughput molecular diagnostics performed on the same bone marrow aspiration. Sequencing was performed in accordance with clinical practices at the Brigham and Women’s Hospital and Dana Farber Cancer Institute, as previously described.^29^ Variants with variant allele fractions (VAF) of greater than 3% were called by default, the large majority of cases in this dataset, with expert review permitting calling of variants with VAFs of <3% (5 cases). Variants of uncertain significance in the *TP53* locus were not included in the dataset. Missense mutations demonstrated VAFs ranging from 0.4% to 95.7%. VAFs of null mutations– defined as frameshift, nonsense, and splice-site mutations– ranged from 3.6% to 93.6%.

We defined disruption of the second allele as a specimen containing a *TP53* mutation with > 50% VAF, detection of at least one additional lower VAF missense mutant, 17p loss by cytogenetics, or absolute or copy-neutral loss of heterozygosity detected by copy number analysis on next generation sequencing.

### p53 mutant residual transcriptional activity scoring

Residual transcriptional activity of p53 mutations was estimated from the Phenotypic Annotation of TP53 Mutations (PHANTM) tool^25^: http://mutantp53.broadinstitute.org/. Wild-type cases were assigned a residual transcriptional activity of 100%, and null cases (frameshift, nonsense, or splice site mutants) were assigned 0% activity. Two cases with missense mutations that were formed by in-frame insertion/deletions, p.H179delinsLN and p.RH213QY were excluded from all analyses that used residual transcriptional activity.

### Statistical analysis and data visualization

Statistical analysis and data visualization were performed using the R statistical computing language with the *data.table*, *psych*, *ROCR*, *magrittr*, *pheatmap*, and *ggplot2* packages. Survival analysis and Kaplan-Meier curves were generated using the *survival* and *survminer* packages. Bootstrapping to compute 95% confidence intervals was performed by sampling 1,000 bootstraps of 80% of the data and using two times the standard deviation of the resulting distribution.

### Code availability

All analysis scripts and raw data to produce figures are available on Github at https://github.com/leeprichman/p53_IHC

## RESULTS

### p53 IHC is specific but not sensitive for detecting *TP53* mutant AML

We retrospectively evaluated 134 bone marrow core biopsies with a pathological diagnosis of AML with p53 immunohistochemistry for comparison to *TP53* mutation status as determined by clinical next-generation sequencing at the Dana Farber Cancer Institute and Brigham and Women’s Hospital. Cases were rated for IHC intensity and percent positivity (see Methods) by raters at three levels of training: anatomic pathology resident, hematopathology fellow, and board-certified attending hematopathologist,. A consensus score of at least 2 of 3 raters was used for diagnostic performance and survival analysis.

As expected, we found a range of ratings intensity for all classes of genotypes (null, wild-type, and missense mutant cases), including 3+ IHC intensity in wild-type AML cases **(Figure 1A)**. Consistent with prior results (**Figure 1B**), p53 IHC was strongly specific (0.93 - 0.97) but only modestly sensitive (0.49 - 0.72) for predicting missense, null, or wild-type *TP53* allele status **(Figure 1C)**, with the highest sensitivity for missense mutations and the lowest sensitivity for null mutations.^12,14–16^

**Figure 1:**
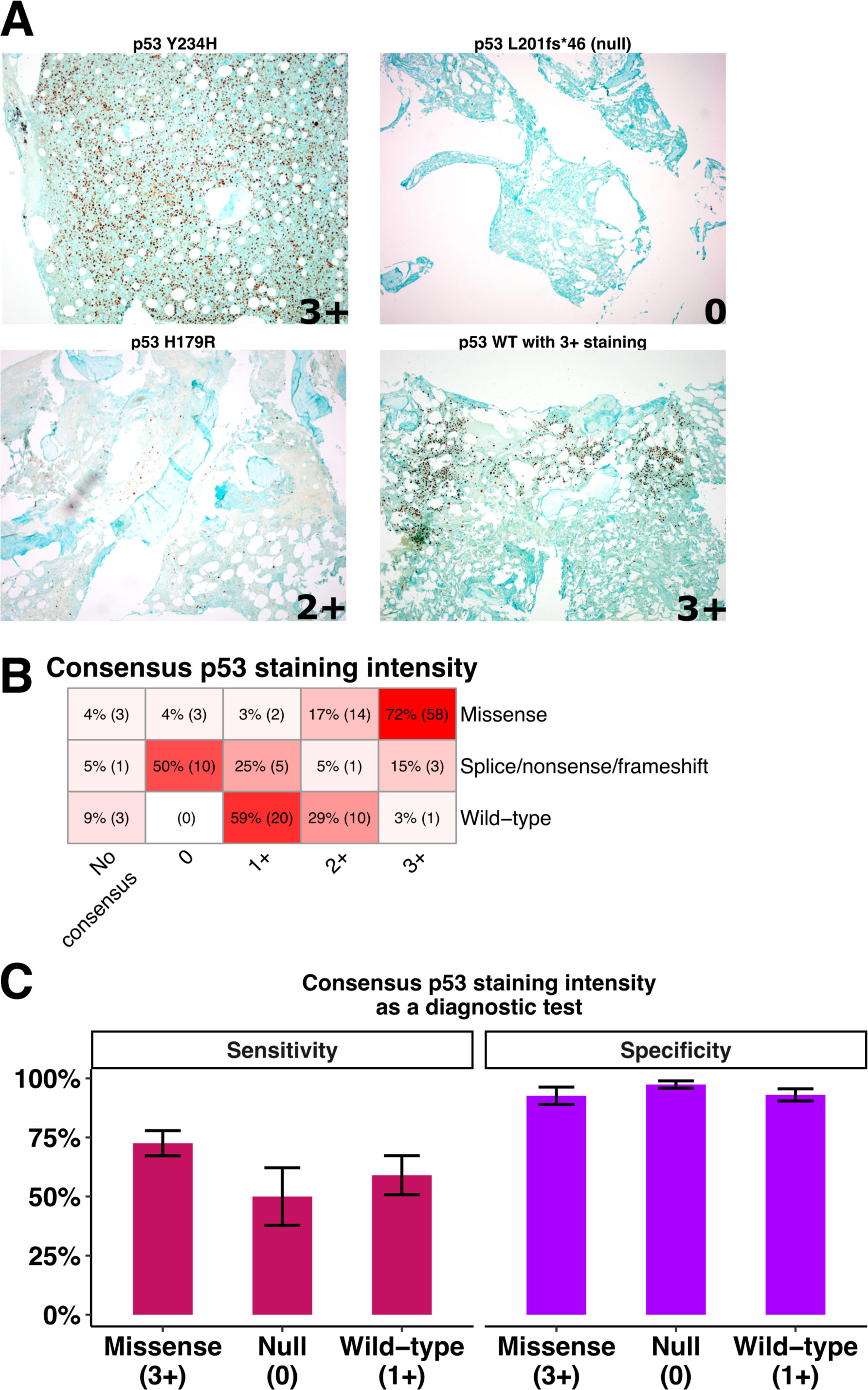
p53 immunohistochemistry (IHC) intensity is variably congruent with mutational status as assessed by sequencing. **(A)** Representative photomicrographs showing p53 IHC staining intensity of 3+ for missense mutant and 0 for null as expected (top), and staining discordant with sequencing results such as ambiguous staining 2+ for a missense mutant, and 3+ staining for a wild-type case (bottom). **(B)** Consensus p53 intensity grading for all cases by *TP53* allele status, percent of cases of that mutation class and absolute number are shown. Consensus score was defined as the IHC intensity with at least 2 of 3 raters in agreement. “No consensus” indicates no raters agreed on the intensity score **(C)** Sensitivity and specificity of consensus IHC intensity to identify missense, null, and wild-type AML. Error bars indicated 95% confidence intervals derived from bootstrapping.

IHC intensity grading had moderate interrater reliability as evaluated by Cohen’s weighted κ (0.74 - 0.91), with expertise-dependence demonstrated by the greatest agreement between the attending hematopathologist and hematopathology fellow. Interrater agreement for 3+ grading was greater for all pairwise comparisons than for 0 intensity grades **(Figure 2A)**. For all pairwise comparisons, percent positivity had statistically significant but weak interrater reliability (Pearson r^2^ = 0.37 - 0.49) **(Figure 2B)**. Together, these findings demonstrate significant deficiencies in the utility of p53 IHC for detecting mutations.

**Figure 2:**
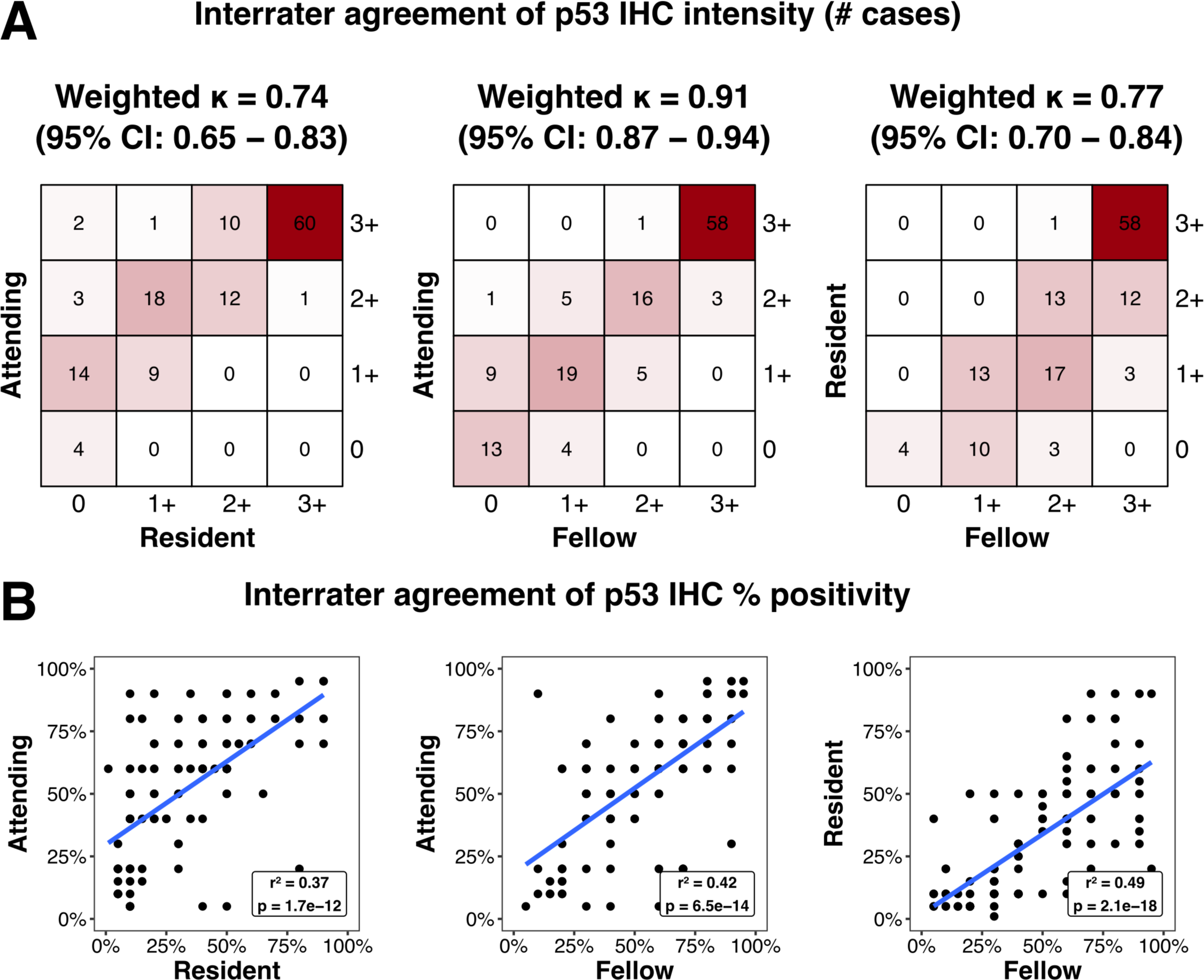
Interrater reliability for p53 staining intensity is modest and expertise-dependent, and overall poor for percent positivity. **(A)** Pairwise comparisons of IHC intensity grading for all raters by level of training by number of cases. Cohen’s weighted κ for interrater reliability is shown. **(B)** Pairwise linear correlation for percent positivity of p53 IHC for raters at all levels of training. Pearson correlation coefficient and associated p-value are shown.

### IHC intensity varies between cases with mutations at identical sites and with identical substitutions

We next systematically evaluated whether cases with mutations at a given locus, whether exhibiting the same or different amino acid substitutions, scored similarly. We identified 19 recurrently-mutated codons in the dataset and found variability in scores for mutations at a given codon and even for identical amino acid substitutions (e.g. P151S, R175H, Y234C, and D281V). We identified instances with identical amino acid substitutions where at least one case had discordant staining from the others **(Figure 3A**). Furthermore, we identified the P151S and R248W mutations as staining with consensus grades of 2+ in the majority of cases, not the expected 3+ of a missense mutant. Additionally, individual cases of G245R and R248Q stained with consensus scores of 1+ and 0, respectively, despite other missense mutations at the same locus staining consistently with the expected 3+ pattern **(Figure 3B)**. Follow-up marrow sampling from individual patients tended to score similarly, although in one instance (cases #44, 45, **Figure 3B**) the consensus scores were discordant (0 vs 3+), suggesting that the same mutation may present differently in an individual over time.

**Figure 3:**
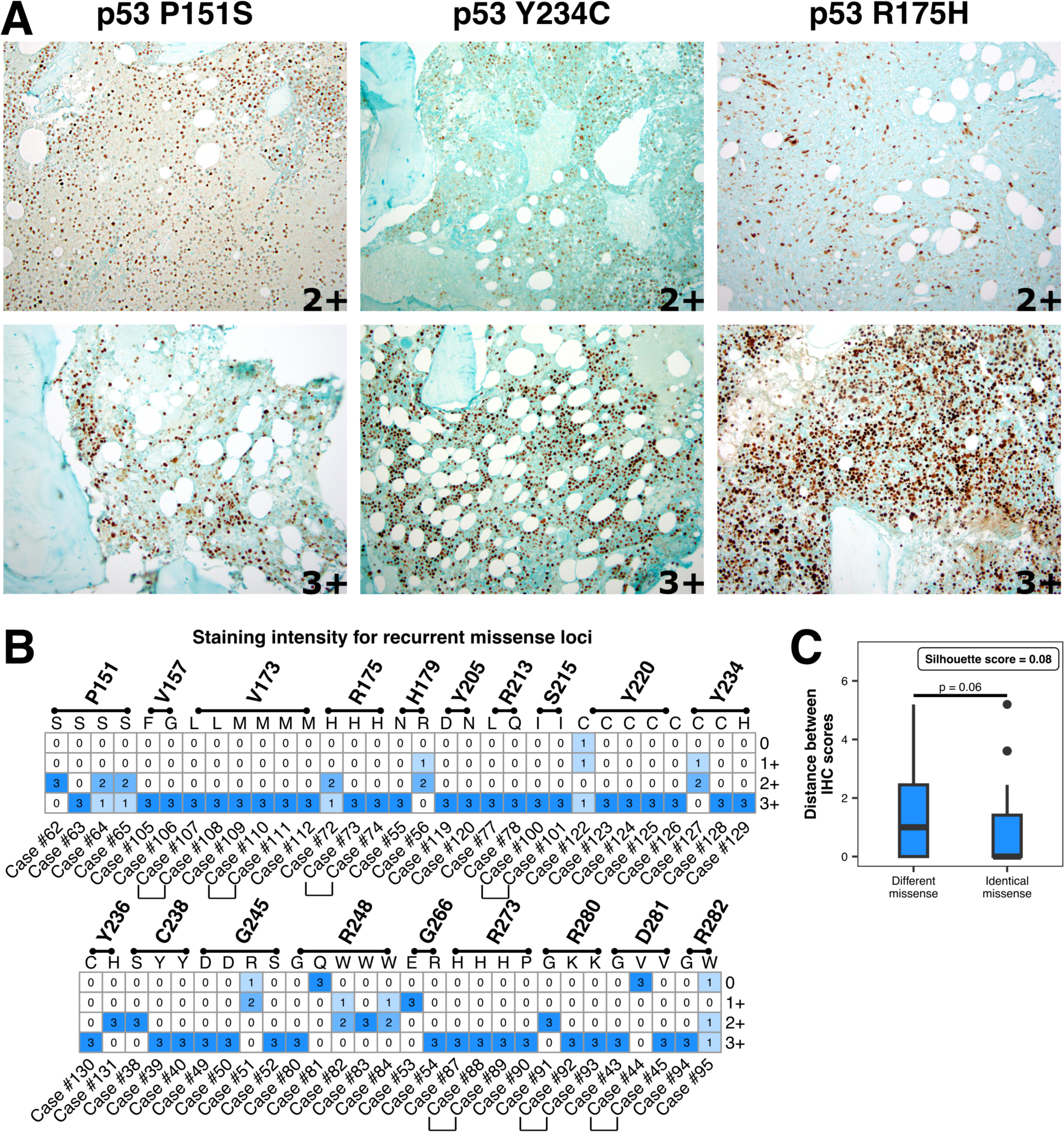
p53 missense mutations have variably consistent staining at the same locus and amino acid substitution. **(A)** Representative photomicrographs of variation in staining intensity across different cases with P151S, Y234C, or R175H mutations. **(B)** Heatmaps showing IHC scores for p53 mutations occurring multiple times in the dataset. Brackets below case numbers indicate samples from different timepoints from the same patient **(C)** Euclidean distance between cases with identical mutations, or between cases with different substitutions at the same locus, were calculated based on IHC scores from 3 reviewers. P-value shown from Wilcox test. Silhouette score, see methods, is shown.

To quantify whether identical mutations, compared to unrelated mutations, were more likely to score similarly across cases, we calculated the distances in IHC scores between cases of the same *TP53* genotype and of different genotypes. Distances spanned similar ranges in both groups, with those between cases of the same genotype tending to have slightly shorter distances, suggesting that their scores were more similar than those of random pairs of cases although this was not statistically significant. We also calculated the silhouette coefficient, which reports on whether a data point is more similar to others in its group (here defined as cases with the same genotype) than to other members of the dataset. A score of 1 indicates that group members are highly similar to each other and highly distinct from other members of the dataset, while a score of −1 indicates the opposite. The silhouette coefficient of 0.08 revealed that, within the limited dynamic range of a 0-3+ IHC intensity scale, cases with identical missense mutations did not have more similar scores than any random pairs of missense mutations **(Figure 3C)**. These findings demonstrate that significant additional variability even within single nucleotide substitutions contributes to the previously demonstrated limited efficacy of p53 IHC^15,16^ for mutation detection.

### Residual transcriptional activity, second allele status, and VAF correlate with 3+ IHC intensity

We then explored other factors that might contribute to discordance between p53 IHC and *TP53* mutation status. Different amino acid substitutions at different sites cause varying levels of dysfunction in the p53 protein and thus affect its transcriptional activity. The phenotypic annotation of TP53 mutations (PHANTM) database contains empirically derived reporter activity for individual amino acid changes throughout the p53 protein.^25^ We interrogated whether this residual transcriptional activity (RTA) of p53 missense mutants could explain whether a missense mutation was rated with 3+ IHC intensity as expected. Receiver operating characteristic area-under-the-curve (AUC) analysis demonstrated that lower residual transcriptional activity (mutations with greater dysfunction) of p53 protein was a weak, but statistically significant classifier of 3+ consensus IHC staining (AUC = 0.60, binomial logistic regression p = 1.4e-5) **(Figure 4A)**.

**Figure 4:**
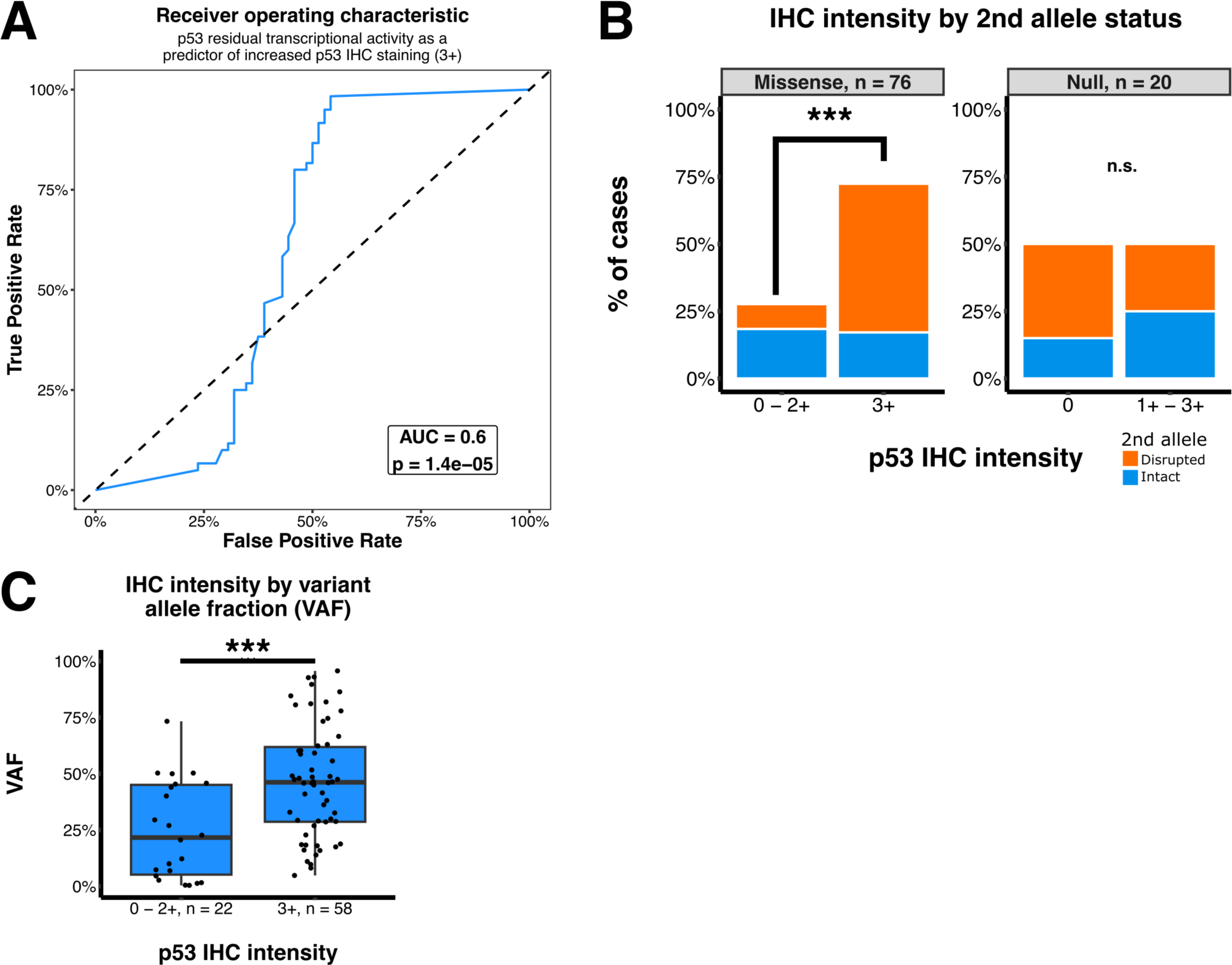
Multiple factors correlate with 3+ p53 IHC intensity of *TP53* missense mutant AML. **(A)** Receiver operating characteristic for residual transcriptional activity of TP53 mutants, retrieved from the PHANTM database, as a predictor of consensus (at least two raters agree) grading 3+ IHC intensity. Area-under-the-curve and binomial logistic regression-derived p-value are shown. **(B)** Consensus staining intensity of missense and null mutants stratified by intactness of the second *TP53* allele determined by next-generation sequencing. P-value shown is for Fisher’s exact test comparing the rate of 2nd allele disruption between the indicated staining intensities. Missense cases without definitive second allele status were excluded (n = 4). “n.s.” = not significant. **(C)** *TP53* variant allele fraction of all p53 missense mutants compared to consensus staining intensity. P-value shown from Wilcox test.

The status of the second p53 allele in these tumors represents another potential confounding factor that influences both mutant and wild-type p53 protein levels (both of which are detected by clinical IHC antibodies). We evaluated whether an intact or disrupted second allele (see Methods) influenced 3+ staining for missense mutants and 0 staining for null mutations. Our analysis demonstrated that missense mutations with 3+ staining by consensus were disproportionately enriched for disruption of the second allele (p <0.001, Fisher’s exact test), which aligns with a previous report.^10^ There was no statistically significant difference in disrupted second alleles amongst null cases that did or did not get rated with the expected zero intensity **(Figure 4B)**. Five cases (#44, 63, 72, 122, and 127) in Fig 3B had discordant staining to other identical mutations. Of those, cases #63, 122, and 127 had intact second alleles which could potentially explain differences observed in IHC intensity. Finally, we identified the VAF of the primary missense mutation as strongly correlating with the tendency to stain with 3+ intensity (p < 0.001, Wilcox test) **(Figure 4C)**. These findings identify unique protein level mutations, the presence of additional hits within the leukemia clone, and the variant allele fraction as factors that influence p53 protein detection by IHC.

### Residual transcriptional activity of *TP53* mutations stratifies overall survival in AML

Previous studies have demonstrated worse treatment response and overall survival for the 10% of myeloid neoplasms with *TP53* mutations.^1–5^ We found that 3+ and 0 p53 IHC intensity both correlated with worse overall survival in our dataset (hazard ratio = 2.4 and 2.1, respectively). As expected, mutant *TP53* allele status determined by sequencing also correlated with worse overall survival for both null and missense mutants (hazard ratio = 4.2 and 2.5, respectively) **(Figure 5A-B)**.

**Figure 5:**
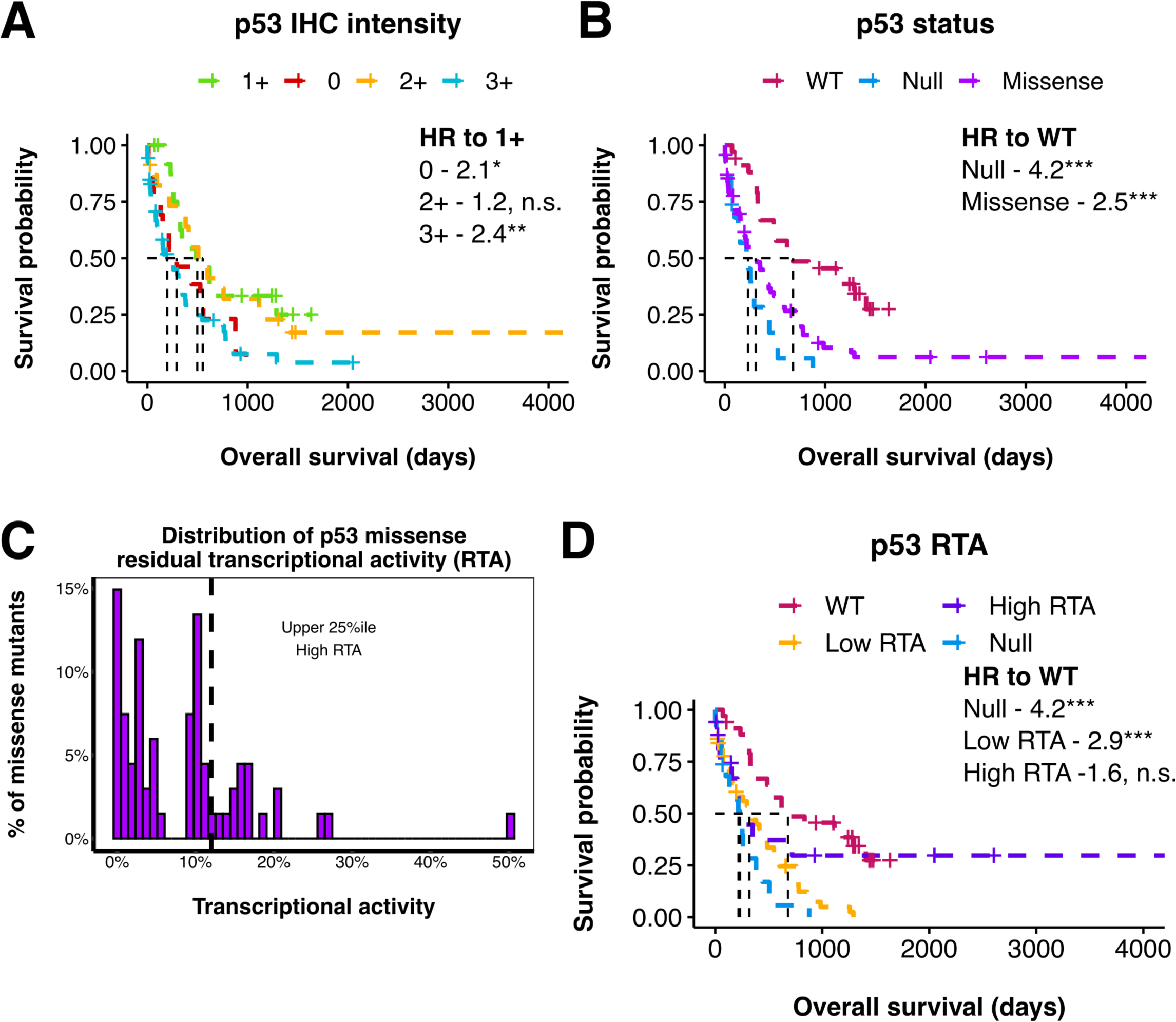
*TP53* mutation confers statistically significantly lower overall survival relative to wild-type AML but not in high residual transcriptional activity mutations. Overall survival of AML cases stratified by consensus IHC staining intensity (at least 2 raters agree) **(A)**, and *TP53* mutation status **(B)**. “WT” indicates TP53 wild-type AML. “Null” indicates frameshift, nonsense, and splice site mutations. Global log-rank p-values were < 0.001 for all comparisons. Hazard ratios (HR) and associated p-values are shown (* - p < 0.05, *** - p < 0.001, n.s - not significant). **(C)** Distribution of TP53 residual transcriptional activity for all AML cases in the dataset, the upper quartile is indicated as “High RTA”. **(D)** Overall survival of AML cases in the upper quartile of residual transcriptional activity (“High RTA”) compared to the lower 75th percentile of residual transcriptional activity (“Low RTA”), wild-type (WT), and null mutations. Hazard ratios (HR) and associated p-values are shown (n.s. - not significant,, * - p < 0.05, ** - p < 0.01).

Given that residual transcriptional activity of mutants correlated with 3+ IHC intensity, we wondered whether mutations with relatively higher intact p53 function correlated with improved prognosis. We compared the upper 25th percentile of p53 residual transcriptional activity in the dataset (>=12.01%) with the lower 75th percentile (“low RTA”), and with wild-type and null mutant overall survival **(Figure 5C)**. Wild-type and high RTA cases did not have a statistically significant difference in overall survival compared to each other. While null mutations and low RTA missense mutations conferred a worse prognosis compared to wild-type (null to WT hazard ratio = 4.2, low RTA to WT hazard ratio = 2.9), there was not a statistically significant difference in hazard ratios between high RTA and null mutation cases **(Figure 5D)**. These findings potentially indicate an intermediate survival phenotype of high RTA mutations compared to p53 null and wild-type AML. We noted that the case with the highest RTA of 50.3% (R181H) did not show the longest survival, suggesting that additional factors, such as comutations, may contribute to the overall survival. Nevertheless, our analyses suggest that individual mutational level analysis, rather than protein detection by p53 IHC, may improve the accuracy of prognostication for *TP53* mutant cases.

## CONCLUSIONS

Rapid assessment of *TP53* mutation status may improve outcomes for patients with this adverse risk factor. While sequencing of the locus is the gold standard, it is not readily available at many centers and can have variable turn-around-times, placing patients at risk of delays in care.^9^ IHC is readily available at smaller facilities, has a rapid turn-around-time, and typically requires no additional expertise to interpret, presenting a compelling alternative for urgent assessment of mutation status.^10–13^ Previous studies have variously reported IHC as inferior,^16,17^ equivalent,^10^ or superior^30^ to sequencing for detecting *TP53* mutations. Most of these studies focused on cases with missense mutations, failing to address the implications of this method for the approximately 20% of p53-mutant AML with inactivating null mutations that have risk for poorer outcomes and lower overall survival in our dataset. Our study found significant deficiencies in IHC as a reliable triage tool for *TP53* status. Interrater reliability was fair, but expertise-dependent. There was heterogeneity in IHC intensity among cases with the same allele status, and sensitivity was moderate overall and poor for null mutations. Compared to previous studies, we found that sensitivity for missense mutations was poorer^31^ or equivalent.^27^ We identified multiple potential confounders that may explain the limited concordance of p53 protein detection by IHC with *TP53* locus sequencing. These deficiencies present a significant challenge for adopting p53 IHC as a rapid surrogate evaluation for mutation status.

Many factors likely contribute to the poor concordance between p53 IHC and *TP53* mutation status identified by sequencing. The unique character of mutations across the *TP53* locus are one source of variability that may influence both the expression levels of mutant protein by IHC and the prognostic implication of such mutations.^4,8,9,13,25,32^ TP53 contains multiple functional domains, each of which may have consequences for protein expression and ultimately clinical outcomes when they harbor a mutation.^2,3,8,18,25,26,32,33^ Even within the DNA-binding domain, the most common site of TP53 mutations in AML, only a few hotspot loci are shared among patients^2,32^, and those which are shared show variability in their IHC intensity in our dataset. Most notably, our finding that even identical mutations may stain differently between bone marrow cores demonstrates additional extragenomic variability in IHC interpretability that presents a significant challenge to clinical validity.

Previous research empirically assessing the residual transcriptional activity of different p53 mutations demonstrates high variability in mutant p53 function between loci and amino acid substitutions.^25^ Wild-type p53 directly drives expression of its own negative regulator, MDM2, which drives p53 degradation.^6,19^ In the missense mutation setting, the absence of functional p53 reduces MDM2 expression, promoting mutant protein accumulation in the nucleus, leading to an increased IHC staining intensity, the so-called “mutant pattern”, seen across cancer types.^20^ Higher residual transcriptional activity of a particular mutant may permit some expression of MDM2 and p53 degradation, which we predict decreases IHC staining intensity. Consistent with this theory, we found that residual transcriptional activity was moderately predictive of 3+ staining. This additional factor of residual transcriptional activity of different missense mutants may partially explain the variability in IHC staining intensity of different p53 mutants and contribute to the discordance with sequencing results.

Human and systematic factors may further confound the interpretation of p53 IHC that contributes to the poor agreement with TP53 sequencing. Sampling error of subclones between marrow cores processed for histology and aspirate samples sent for sequencing may add additional variability.^21,22^ The abundance of blasts and variant allele fraction of mutant p53 alleles may further confound interpretation of IHC, as rare 3+ staining may be missed in a low tumor burden sample.^21^ Conditions of tissue fixation and decalcification in bone marrow cores are another potential source of technical variation. Furthermore, strong nuclear expression in erythroid progenitors, often increased under the anemic conditions of marrow involving disease or cytotoxic chemotherapy, also impedes accurate interpretation.^23,34^ Together, these factors introduce a largely uncontrollable source of variation that reduces the concordance between IHC staining patterns and *TP53* mutation status.

UItimately, our study found that p53 IHC is a limited surrogate measure for *TP53* mutation status. While IHC intensity was specific for mutational status and correlated with overall survival, it had limited sensitivity. Multiple sources of phenotypic and technical variability likely limits sensitivity and confounds concordance between p53 protein expression as detected by IHC and gold standard sequencing results. The residual transcriptional activity of p53 mutants represents an underexplored factor that may influence IHC and other diagnostic studies and clinical outcomes and merits further exploration.The modest sensitivity of p53 IHC for detecting cases at risk for adverse prognosis limits its use as a screening tool. Development of low-resource, rapid sequencing-based methods of TP53 mutation analysis may yield a more reliable clinical triage tool.

## Data Availability

All data produced are available online at github.com/leeprichman/p53_IHC

## ACKNOWLEDGEMENTS

The authors would like to thank Dr. Elizabeth Morgan, Dr. Sarah Wu, Dr. Gabriel Griffin, Dr. Nagarajan Nandagopal, and Meg Dillingham-McCullough for their helpful commentary on the manuscript. We would also like to thank Dr. Geraldine Pinkus, Alyson Smeedy Campbell, and the staff of the histology and IHC labs in the Department of Pathology at Brigham and Women’s Hospital. This work was supported by the Ludwig Center at Harvard (AJ).

